# Approaches for measuring socioeconomic status in health studies in Sub-Saharan Africa: a scoping review

**DOI:** 10.1101/2025.01.01.25319868

**Authors:** Daniele Sandra Yopa, Gbetogo Maxime Kiki, Patrice Ngangue, Marie Nicole Ngoufack, Gilles Protais Lekelem Dongmo, Douglas Mbang Massom, Anya Amvella Priscillia, Brian Bongwong Tamfon, Alain Chichom-Mefire, Catherine Juillard, Alan Hubbard, Georges Nguefack-Tsague

## Abstract

**Background:** Socioeconomic status (SES) is essential for determining a person or community’s position about certain social and economic characteristics. This is particularly important in sub-Saharan Africa, where health disparities are pronounced. We conducted a scoping review to explore approaches used in health studies to measure socio-economic status in the sub-Saharan region.

**Methods:** A comprehensive literature search covering January 2012 to June 2024 was conducted in five databases: PubMed, EMBASE, CIHNAL, Web of Science, and African Index Medicus. All studies in sub-Saharan Africa focused on health-related socioeconomic status were included, regardless of study methodology. Three peer reviewers independently evaluated the selected articles according to inclusion and exclusion criteria. Discrepancies between reviewers were resolved through a consensus meeting. The review protocol was registered on the Open Science Framework (OSF, <u>OSF.IO/7NGX3</u>).

**Results:** The initial search yielded 19,669 articles. At the end of the screening process, 65 articles were analysed. Cross-sectional studies have been widely used. South Africa (13.4%) and Kenya (11%) were the most represented countries. Maternal, neonatal, and infant/juvenile health was the most covered theme (31%). The review identified 12 categories of SES measurement methods, with the asset-based wealth index being the most widespread (61.9%). Principal component analysis (PCA) is the primary analytical method used to calculate this index (57.7%).

**Conclusions:** This scoping review identified the asset-based wealth index as the most frequently used and provided essential elements for pooling different SES calculation methodologies to reach a consensus. Using SES to improve interventions is important to limit African health disparities.

## 1. Introduction

Socio-economic status (SES) is crucial in explaining health disparities, more precisely in access to care and health coverage in vulnerable communities [1–3]. It is also recognized as an essential determinant for improving public health policy and is important to track progress towards Sustainable Development Goals (SDG) related to health [4–6]. Its calculation generally integrates several components, such as the standard of living, assets, and economic, social, and professional life [7]. At the dawn of the emergence and reemergence of infectious and chronic non-communicable diseases in Africa, SES allows the prioritization of interventions in at-risk areas and populations and measures their impacts on improving health equity [8–11]. However, the calculation approaches used to grade the SES vary according to the indicators used and can strongly influence the recommendations made by stakeholders.

Due to the diversity of socioeconomic contexts and conditions, measuring SES is complex. In addition, the indicators usually used differ according to the SES calculation method and expected objectives. Moreover, the indicators used to assess SES may vary depending on the population studied and the availability of data [12]. However, this variability makes it difficult to compare studies because of the diversity of realities in countries and communities [11, 13–14] Finally, developing a standardized and consensual approach across Africa is a significant challenge because of fluctuations in the accuracy and credibility of the measures used [15–17].

Health inequalities are a significant problem with enormous consequences for human capital. African countries, which have a heavy burden of infectious and non-communicable diseases, need to understand the socio-economic determinants of health to guide health interventions efficiently [18]. Sub-Saharan Africa is characterized by a disparate socioeconomic context due to the political structure and cultural and historical legacies that must be considered in defining and selecting indicators for measuring [18,19]. However, the lack of standardized SES measures can hamper efforts to tackle the wide range of inequalities in high-risk groups and the assessment of interventions implemented to curb them. Given the above challenges, this scoping review aims to provide a comprehensive overview of the different approaches used to measure SES in sub-Saharan African health research. We explored the following research questions: How is socioeconomic status (SES) measured in health studies and in terms of well-being in Africa? What are the variables and methodologies used to determine SES? It will probably improve the understanding of SES measurement while providing valuable information that can inform future studies, improve public health interventions, and reduce health disparities across the continent.

## 2. Methods

This scoping review used literature searches to address broad research questions, incorporate data from available quantitative and qualitative methodologies, and summarize the main findings [20]. Our review followed the methodological framework developed by Arksey and O’Malley [21]. We included relevant literature, regardless of the study design or evidence quality. Our scoping review protocol was registered on the Open Science Framework (OSF) at the following link: https://doi.org/10.17605/OSF.IO/7NGX3.

### 2.1 Search strategy

The following databases were searched to identify relevant publications: PubMed/Medline, CINAHL, EMBASE, Web of Science, and African Index Medicus from January 2012 to June 2024. The key search terms for the population, intervention, and outcomes are listed in Table 1. We choose the latest 12 years to ensure that the conceptualization of SES reflects contemporary indicators. We included all types of manuscripts, guideline reports or editorials published in English, focusing on SES and African countries. Conference proceedings, books, grey literature, case reports, letters, notes, and studies on “non-Sub-Saharan” African countries were excluded.

**Table 1:** Keywords used for developing a comprehensive search strategy.

| Population (P) | Intervention (I) | Outcome (C) | Filter |
| --- | --- | --- | --- |
| ✓ Family | ✓ Theoretical model | Health | All the countries in Sub-Saharan Africa |
| ✓ Population | ✓ Economic model |  |  |
| ✓ Residence | ✓ Socioeconomic factors |  |  |
| ✓ Family characteristics | ✓ Social class |  |  |
|  | ✓ Socioeconomic Status |  |  |
|  | ✓ Socioeconomic disparity |  |  |
|  | ✓ Wealth index |  |  |

### 2.2 Screening and Selection

The Rayyan platform was used to compile and screen articles. Duplicates were removed. A team of reviewers (SYD, GMK, PN, MNN, DMM, LDGP, AAP) conducted the selection process in two stages following the previously stated inclusion and exclusion criteria. A couple of reviewers independently evaluated each article. In the first stage, selection was made based on the titles and abstracts. The second stage involved full-text screening. Each reviewer documented the reasons for exclusion on the platform. Disagreements were resolved by consensus during a plenary meeting involving all the reviewers. Each selection phase began with a benchmarking exercise to minimize errors and reduce the risk of discrepancies between the reviewers.

### 2.3 Data extraction

Relevant information for each study was extracted using a pre-developed grid. The extracted data included details, such as the first author’s name, article title, type of article, objectives, study design, sample size, study period, data source, population profile, SES measure name, countries involved, methodology, and indicators used. SES measures were analyzed and categorized based on the calculation methods and types of indicators used.

## 3. Results

The database search yielded 19,669 publications. After removing duplicate articles; 15,215 articles remained for title and abstract screening. Of these, 15,029 which did not meet the inclusion criteria were excluded. Of the 186 articles selected for full-text screening, 15 were excluded because full texts were unavailable. Ultimately, 65 articles were included in the final analysis. Figure 1 provides details of the search process (see Figure 1).

**Figure 1:**
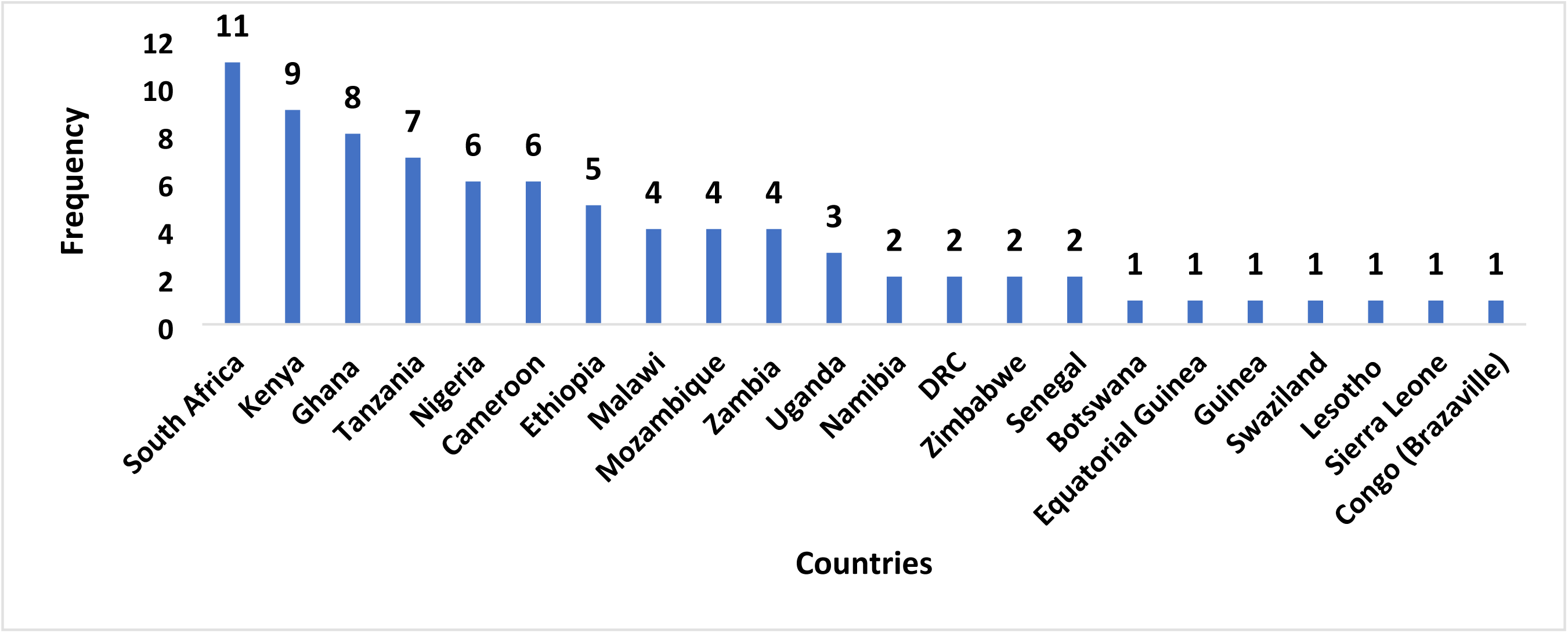
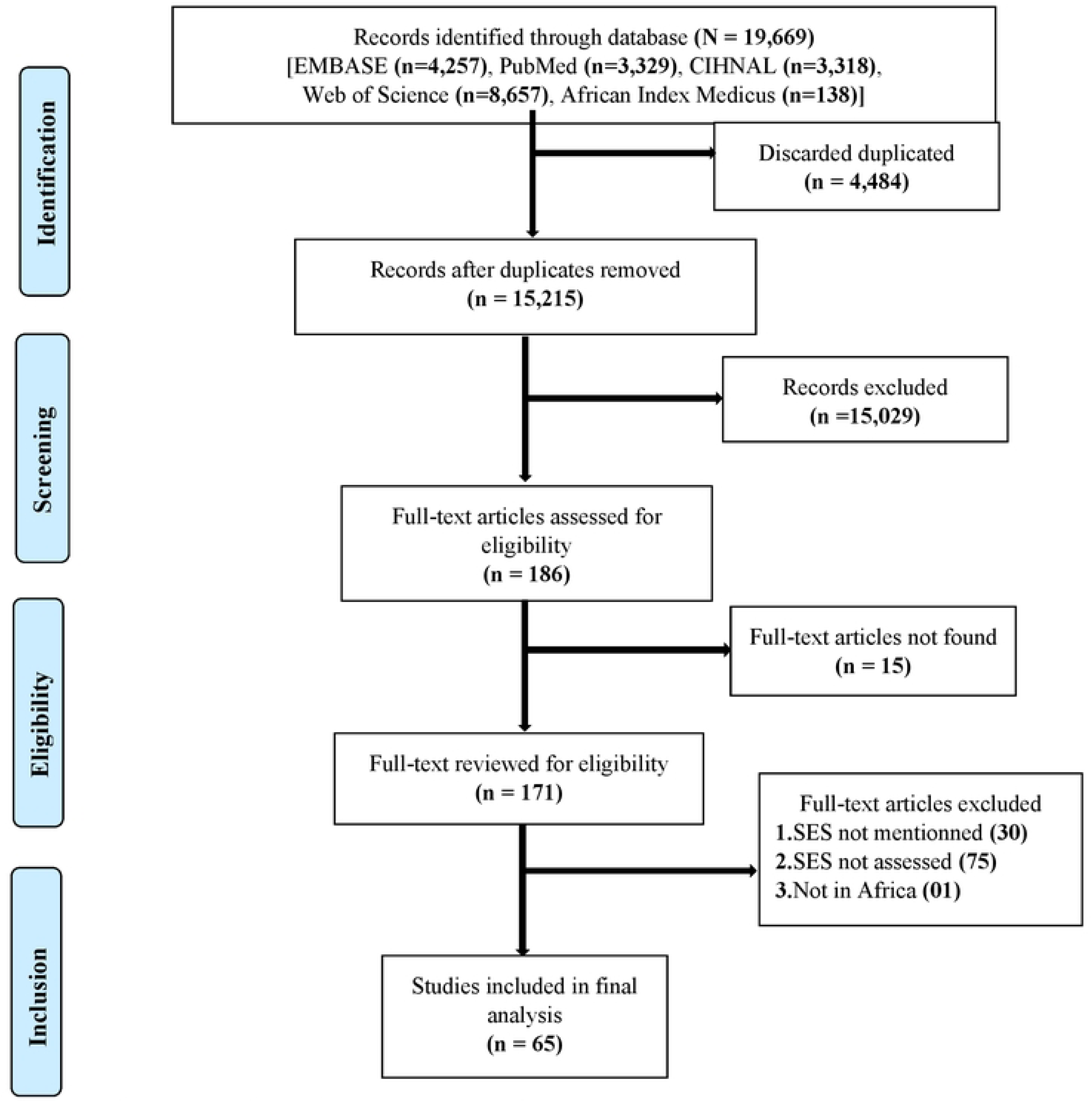
Frequency of mentioned Countries in the articles of Scoping review, n= 82

### 3.1 Study Characteristics

#### Geographical distribution of the studies

A total of 22 countries were included in this review. Figure 2 shows that the most frequently referenced articles were from South Africa (13.4%, n= 11 articles), Kenya (11%, n= 9), and Ghana (9.8%, n = 8).

**Figure 2:**
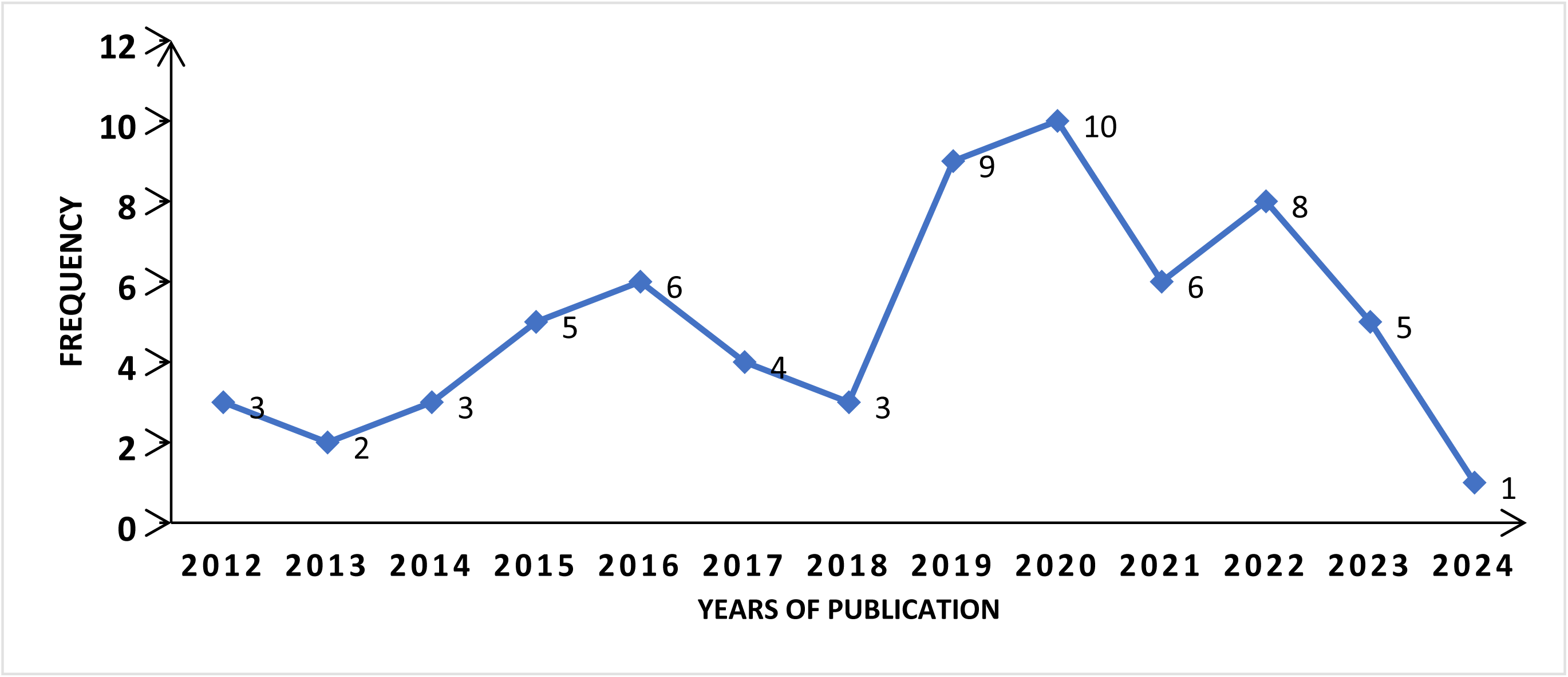
Number of articles published per year, n= 65

#### Number of articles published per year

Figure 3 shows that the highest number of publications was published in 2020, with 15% (n=10 articles), and 2022 with 12% (n=8).

#### Characteristics of included studies based on the type of article, design, and field of intervention

The research article was the most common type of article, accounting for 93.8% (n= 61). The study design most represented was a cross-sectional study (52%, n=34), followed by cohort studies (23%, n= 15). Socioeconomic status and maternal and child health were the most frequently covered themes, representing 31% each. The sample size ranged from 333 participants to 1,273,644 participants.

### 3.2 Socioeconomic status measures

#### Different category of SES Indices and measurements

Eleven SES categories with 41 measures of socioeconomic status were identified in the 65 selected articles. The most used method for socioeconomic status calculation was the asset-based Wealth Index (Table 3), with 61.9% (n = 52), with the Wealth Index (WI) representing 28.6% (n= 24). The WI includes variations such as the New WI, Household WI, Harmonized Wealth Index, and Absolute WI. Table 2 outlines various indices and their calculation characteristics.

**Table 2:** Characteristics of included studies, n= 65.

| **Characteristics of included studies** | **Number of articles**<br>**(n= 65)** | **Percentage**<br>**(%)** |
| --- | --- | --- |
| **Type of article** |  |  |
| Research article | 61 | 93.8 |
| Methodology article | 4 | 6.2 |
| **Study design** |  |  |
| Cross-sectional study | 34 | 52 |
| Cohort study | 15 | 23 |
| Quantitative data analysis | 8 | 12 |
| Mixed methods involving qualitative and quantitative approaches | 3 | 5 |
| Case-control study | 2 | 3 |
| Experimental/Quasi-experimental study | 1 | 2 |
| Methodological study | 1 | 2 |
| Review | 1 | 2 |
| **Field of intervention** |  |  |
| Socioeconomic status assessment, wealth, poverty | 20 | 31 |
| Maternal, Child, youth, family health and wellbeing | 20 | 31 |
| Non-communicable diseases (NCDs) and control (including trauma and eye health) | 8 | 12 |
| Neglected tropical disease control and malaria | 4 | 6 |
| Mental health | 3 | 5 |
| Communicable disease (TB, HIV) | 2 | 3 |
| Nutrition | 2 | 3 |
| Gerontology | 2 | 3 |
| Public health research and program evaluation | 2 | 3 |
| Social determinants of health | 2 | 3 |
| **Total** | **65** | **100** |

**Table 3:** Different categories of socioeconomic status indices found in the scoping review, n= 84.

| Categories | Included articles | Frequency<br>(n= 84) | Percentage<br>(%) |
| --- | --- | --- | --- |
| **SES Measures** |  |  |  |
| **Asset-based wealth Index** |  |  |  |
| Wealth Index | [22-45] | 24 | 52 (61.9) |
| Household SES | [46-48] | 3 |  |
| Asset Index calculation | [49-50] | 2 |  |
| Socioeconomic Position | [49,51] | 2 |  |
| Socioeconomic Status | [52, 53] | 2 |  |
| Global Network Socioeconomic Status Index (GN-SESI) | [54, 55] | 2 |  |
| Utility-based living standards index (ULS) | [56] | 1 |  |
| Childhood SES Index | [38] | 1 |  |
| Neighbourhood SES Index | [46] | 1 |  |
| Socioeconomic Index | [57] | 1 |  |
| PCA Index | [31] | 1 |  |
| SHINE Wealth Index | [58] | 1 |  |
| Polychoric Dual-Component Wealth Index (P2C) | [59] | 1 |  |
| Objective relative wealth | [60] | 1 |  |
| Multidimensional wellbeing indicator | [61] | 1 |  |
| Economic Clusters model | [16] | 1 |  |
| Asset indices (MCA index) | [31] | 1 |  |
| Household SES using the MCA model | [62] | 1 |  |
| Socioeconomic Status (SES) indice MCA model | [63] | 1 |  |
| Wealth quintiles (EquityTool) MCA model | [64] | 1 |  |
| Household Wealth Index with RF | [42] | 1 |  |
| DHS Wealth Index (WI) with RF | [65] | 1 |  |
| Census Wealth Index (CWI) with RF + geographical location | [65] | 1 |  |
| **Indices based on Education, health, and Living standards** |  |  |  |
| Multidimensional Poverty Index | [39,42,50,66-74] | 13 | 14(16.7) |
| Fuzzy poverty Index | [75] | 1 |  |
| **Index based on Occupation, Income, Education** |  |  |  |
| Wealth asset, Education, Occupation (WEO) | [42] | 1 | 4(4.8) |
| Household SES in Cameroon (occupation, income, educational level) | [76] | 1 |  |
| Individual SES index | [77] | 1 |  |
| Objective Social Status (OSS) | [78] | 1 |  |
| **Index based on perceived wealth, Income, Education, and Occupation** |  |  |  |
| Subjective Social Status (SSS) | [78,79] | 2 | 4(4.8) |
| Subjective SES index (SSES) | [77] | 1 |  |
| Subjective relative wealth (SRW) | [60] | 1 |  |
| **Asset based wealth indices with material affluence scale** |  |  |  |
| SES using Material Affluence Scale (MAS) | [80,81] | 2 | 2(2.4) |
| **Index based on Income** |  |  |  |
| Poverty score (Simple Poverty Scorecard) | [64] | 1 | 2(2.4) |
| Social status | [41] | 1 |  |
| **Index based on education, occupation** |  |  |  |
| Individual-level SES (Educational attainment and occupational status) | [82] | 1 | 1(1.2) |
| **Index based on asset, income, disability** |  |  |  |
| NHIF scorecard | [25] | 1 | 1(1.2) |
| **education and Income** |  |  |  |
| WAMI | [42] | 1 | 1(1.2) |
| **Asset wealth Index based + income** |  |  |  |
| Household SES index | [77] | 1 | 1(1.2) |
| **Asset based on Expenditure** |  |  |  |
| Household economic status | [83] |  | 1(1.2) |
| **No name** |  |  |  |
| S.E.S (no explicit name) | [84] | 1 | 1(1.2) |

#### Indicators and methodology analysis used by category of SES Index

This section presents the indicators for calculating socioeconomic status for the most common indices found (see Table 4).

**Table 4:**
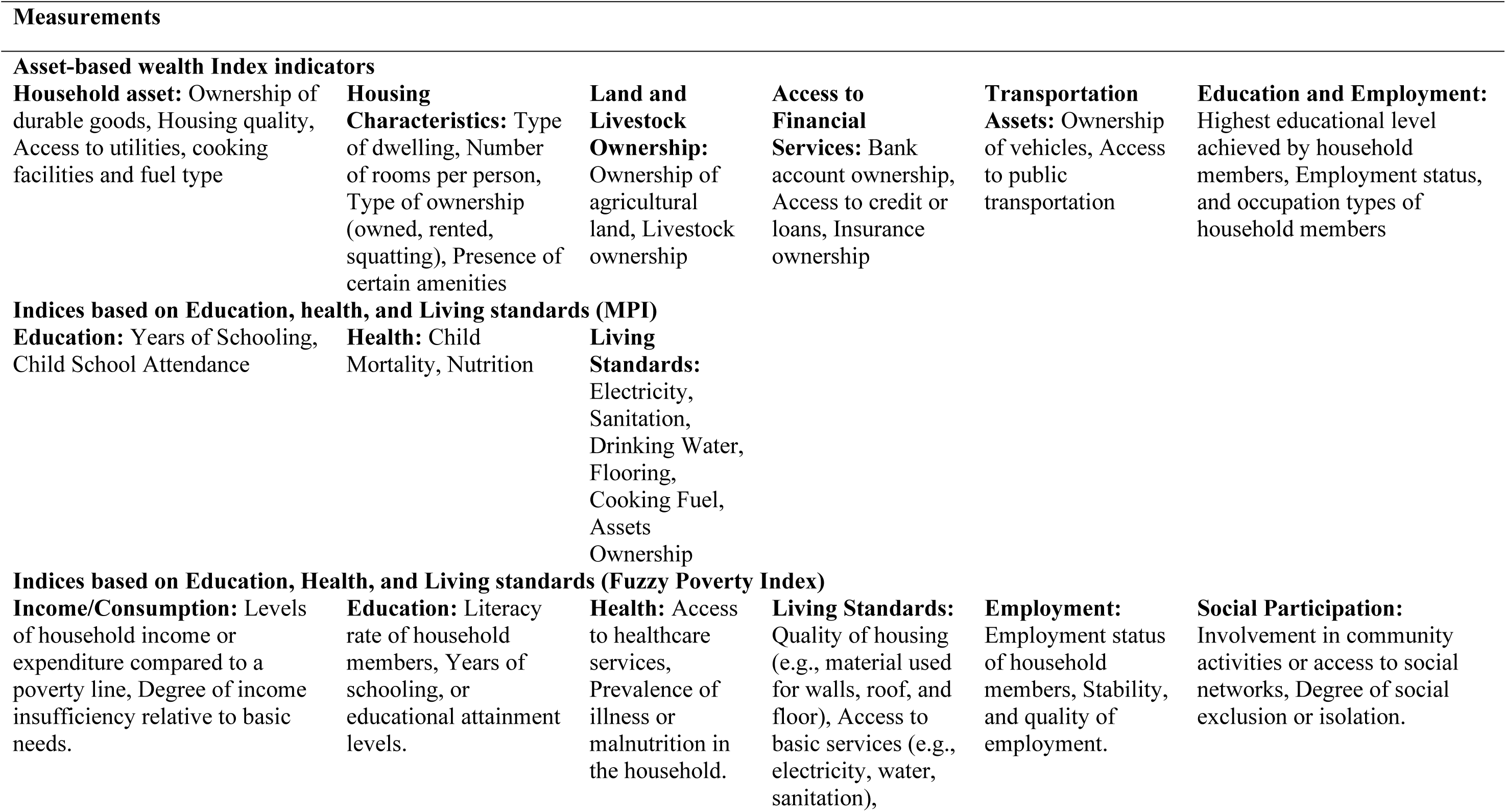
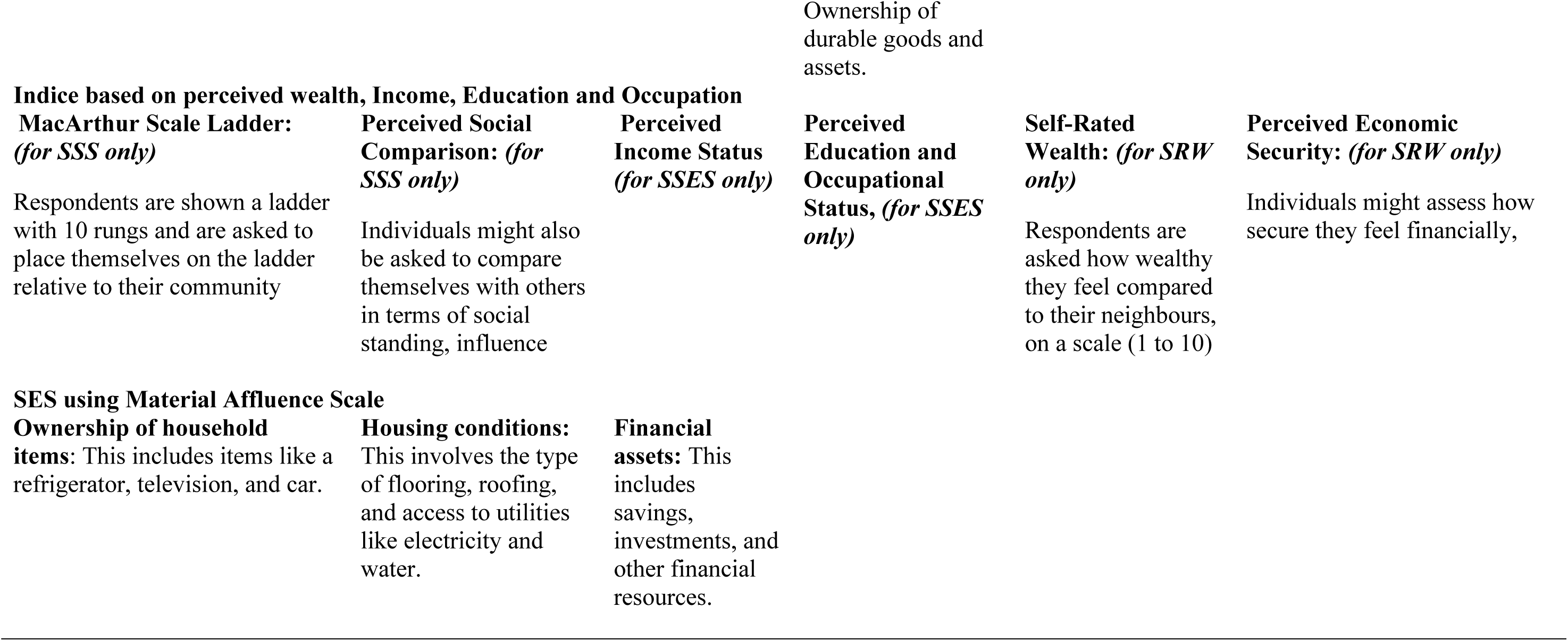
Common indicators used by the most frequent category of the SES Index.

Table 5 displays the analysis methods most frequently used in the Asset-Based Wealth Index. PCA was the predominant method, used independently in 57.7% of cases and in combination with other methods in 9.6% of cases.

**Table 5:**
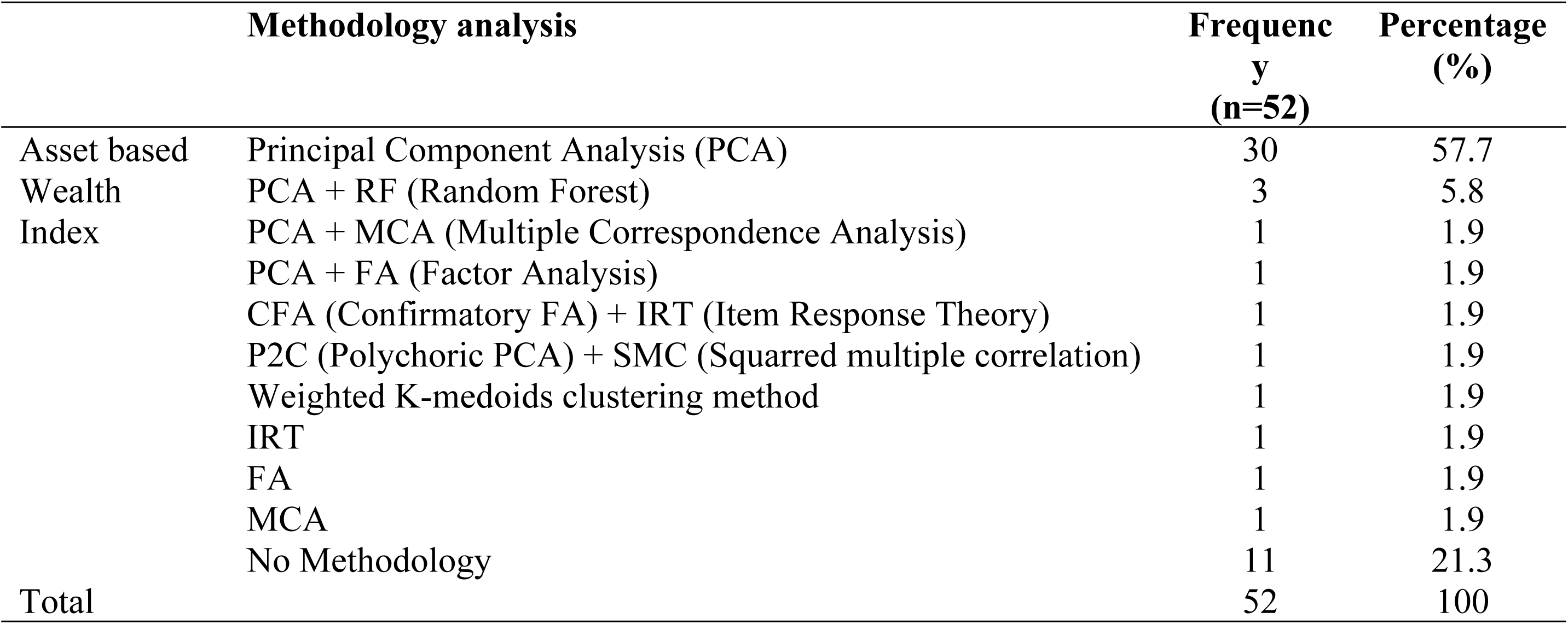
Methodology analysis used to calculate Asset Based Wealth Index.

### 1.4. Discussion

This study aimed to explore methods for measuring SES in African health studies. After rigorous screening, 65 articles were included in the final analysis, providing a robust dataset for examining various aspects of health disparities. This review revealed 41 different types of SES measures, which can be categorized into 11 groups. The asset-based wealth index is the most used SES measure, followed by the indice-based wealth index based on income, education, and living standards. South Africa, Kenya, and Ghana are the most representative countries. This distribution may reflect differences in data availability in some countries. The increase in the number of publications over time, especially between 2020 and 2022, suggests a growing interest in SES measurement in African health studies exacerbated by the COVID-19 pandemic. This trend is consistent with the critical review by Dagher and Linares, which highlights the complex interaction between the social determinants of health and adverse health outcomes, particularly during crises [2].

The prevalence of cross-sectional studies (52%) and cohort studies (23%) indicates an interest in identifying the determinants of short- and long-term health disparities, as demonstrated by the work on economic status assessment in Eyler and Hubbard trauma registries [85]. The particular interest in the theme of maternal and child health demonstrates that efforts are being made to understand disparities within vulnerable groups, as described in the studies by Adler et al. and Alamneh et al. in the field of maternal and child health [1,10]. To achieve the Sustainable Development Goals 3 aimed to reduce maternal mortality to less than 70 per 100000 live births and mortality to less than 12 per 1000 live births by 2030, efforts should be made to enhance the coverage of essential and adequate maternal and child healthcare services [86].

A wide variety of methods have been used to measure SES. Of the 41 indices identified, the wealth by asset index was the most used, accounting for 61.9% of all measures. This index, often calculated using principal component analysis (PCA), considers elements such as possession of durable goods and quality of housing and defines long-term wealth [87]. It uses data from demographic health surveys with a standardized data collection tool in several countries, including those in Africa. This facilitates comparability among countries with limited resources [88]. The Wealth Index (WI) and its variations, such as the New WI, Household WI, Harmonized Wealth Index, and Absolute WI, were used frequently, accounting for 28.6% of the indices. The variability of the WE calculation indicates a continuous effort to adapt, to better understand the nuances of economic conditions in different contexts. The use of multiple indices follows the work of Batool and Hennig, who proposed clustering methods to improve the accuracy of socio-economic assessments [89].

The main component analysis (PCA) appeared to be the predominant method for asset-based wealth index calculation, used independently in 57.7% of cases and in combination with other methods in 9.6% of cases. The dependence on PCA is due to its ability to synthesize several variables together when they are all quantitative to best describe the set of individuals defined by these variables in the descriptive study. This reduces the number of initial variables while returning the maximum amount of information [90,91]. The use of other methods, such as Multiple Correspondence Analysis (MCA), could be an alternative method of calculating socioeconomic status in households [87]. Studies have shown that combining PCA and MCA can be beneficial, with neither method used to the detriment of the other, but rather to complement it [92].

Different SES measurement methods can significantly impact the detection and interpretation of health disparities. In the case of less used measures, such as SSS, which allow to understand how different groups of people perceive themselves in the social hierarchy, their unique interpretation among groups can maximize positive health outcomes [93].

Indices based on income and expenditure are more direct measures of short-term socioeconomic status, but data are often difficult to collect from households in developing regions, where informal employment remains significant [94]. Asset-based indices seem to be more accurate in identifying disadvantaged groups in contexts where income is unreliable. Policy decisions based on unreliable indices could worsen equity, meaning that resources might not be allocated fairly or effectively [95]. Studies that use multidimensional measures, such as MPI, tend to reveal more pronounced health disparities, particularly about infant mortality, malnutrition, and access to healthcare.

The results of this study highlight the importance of choosing an appropriate SES measure according to the context and theme addressed. Policies based on simplistic SES measures such as income alone may not be effective in targeting vulnerable groups. Some healthcare subsidy programs that do not consider certain indicators may not reach those who need it most.

## Strengths and Limitations

The challenge of drawing strong conclusions on a continental scale lies in the diverse methods used to calculate SES, from asset-based indices to those that incorporate dimensions, such as education, income, perceived wealth, and health.

Regarding the study’s limitations, it should be pointed out that, like many scoping reviews, we did not conduct a thorough critical appraisal of the quality of the studies included. Additionally, the comparability of the studies due to the heterogeneity of methodologies and approaches is another limitation to consider. Lastly, we acknowledge that we may not have thoroughly reviewed all of the available literature on our topic.

## Conclusion

The purpose of this review was to map all available indices and methods used to measure socio- economic status in health-related studies in sub-Saharan Africa. Principal Component Analysis is the most used calculation method. The asset-based wealth index was the most widely used, followed by indices focusing on education, health, living standards, and many other indices with their specifications. These measures can be utilized by researchers to provide valuable information that informs policies and intervention strategies that aim to reduce health disparities and promote equity. It is recommended to move towards the standardization of SES measurement methods in Africa, while allowing adjustments for local contexts.

## Data Availability

It is a scoping review, thus no data available

## Acknowledgements

None

